# Effect of Monitoring and Evaluation Systems on the Performance of Neonatal Intensive Care Unit at Yumbe Regional referral hospital; A Pre-post quasi-experimental study design

**DOI:** 10.1101/2024.09.16.24313784

**Authors:** Innocent Ssemanda, Patrick E. Odong, Mubaraka Nasur, David Ejalu, Karen Mwengwe, JMO Tukei

## Abstract

**Purpose:** This study explored the effect of implementing monitoring and Evaluation (M&E) systems on the performance of the Neonatal Intensive care Unit at Yumbe regional referral hospital.

**Methods:** A pretest-posttest quasi-experimental design was employed involving 236 neonates, with 103 in the pretest group (Before the implementation monitoring and evaluation systems) and 130 in the posttest group (after the implementation of monitoring and evaluation). The intervention lasted 105 days. Key performance indicators (KPIs) such as; neonatal mortality rates, length of stay, neonatal intensive care’s effectiveness, morbidity rate, survival rates, and infection control were measured. Patient satisfaction as a secondary outcome was also explored through questionnaire surveys. Data collected was entered directly in Micro software, and exported to the STATA version 18 for analysis

**Results:** Neonatal mortality rates significantly decreased from 19.4% in the pretest group to 7.7% in the posttest group (P<0.01). The survival rates improved from 80.6% in the pretest group to 92.3% in the posttest group. The average length of stay was reduced from 10 days (SD=4) to 8 days (SD=3) (P<0.05). Neonatal intensive care effectiveness scores improved from a mean of 2.8 to 3.5 (P<0.01). Compliance with Neonatal intensive care unit protocols increased from 70% to 80% (P<0.01). The reliability of monitoring and evaluation components was high, with Cronbach’s alpha values ranging from 0.754 to 0.915

**Conclusion:** Implementation of monitoring and evaluation systems significantly enhanced NICU’s performance, reduced mortality rate, improved survival rates and improved patient satisfaction. These findings underline the importance of M&E frameworks in optimizing neonatal care.

## Introduction

Neonatal mortality remains a critical public health challenge globally, with Uganda being no exception. Despite advancements in healthcare systems, preventable newborn deaths continue to occur at alarming rates in many Ugandan hospitals (Egesa et al., 2020; Tibaijuka et al., 2021). This persistent issue highlights the need for effective interventions to improve neonatal care outcomes. One promising approach is the implementation of robust Monitoring and Evaluation(M&E) systems (Kevany et al., 2012; Ogungbemi et al., 2012), within the Neonatal intensive care unit (NICU). These systems are hypothesized to enhance the quality of care, streamline neonatal clinical care processes, and ultimately reduce neonatal mortality rates, by identifying performance gaps in healthcare service delivery, and facilitating timely interventions(Mohamud, 2023; I. M. Njeru & Luketero, 2018). By systematically tracking and analyzing NICU’s key performance indicators, M&E systems can provide actionable insights that can lead to improved clinical practices and better health outcomes for newborns in Uganda(Apondi, 2023). Unfortunately, this model is not yet adopted in many NICU facilities. Furthermore, the incorporation of M&E systems components into NICU’s routine activities is lacking, and this due to a lack of evidence to suggest that, the performance of the Neonatal Intensive Care Unit (NICU) improves when integrated monitoring and evaluation(M&E) systems. This fact remains underexplored, raising concerns about meeting the World Health Organization’s target of 12 neonatal deaths per 1,000 live births by 2030 (U. WHO, 2020). Back centuries, the lack of advanced NICU technology hindered efforts to save critically ill neonates (Budetti & McManus, 1982; Glazebrook et al., 2007; Horbar & Lucey, 1995). Gratitude to Louis Gluck for introducing modern NICU technology in 1960s which marked a significant advancement in providing specialized care to premature and low-birth-weight infants, and enhanced survival rates(Gartner, Gartner, Gluck, & Butterfield, 1992). Despite these advancements, recent evidence suggests a decline in NICU’s performance, complicating efforts to achieve Zero neonatal mortality by 2030 (Sharrow et al., 2022). The current literature further reports high neonatal mortality rates, and reduced survival rates among sick newborns admitted to the NICU, and this has been linked to poor health information systems management and a lack of M&E system mechanisms (Getabelew, Aman, Fantaye, & Yeheyis, 2018; Woday Tadesse, Mekuria Negussie, & Aychiluhm, 2021).

Globally, neonatal mortality rates remain high, with an estimated 2.4 million children dying in the first 28 days of months, of their life in 2019, and Africa contributing a significant 60-percentage increase (Unicef, 2021a; WHO, 2021). In Uganda, neonatal mortality rates are alarmingly high, with over 40.758 deaths per 1, 000 live births in 2021, and 27 deaths per 1,000 live births in Yumbe-West Nile region births (UN, 2021). Efforts are put in place to reduce these rates including the introduction of; maternal and prenatal death reviews and surveillance (MPDRS), weekly death surveillance, Kangaroo mother care, Exclusive breastfeeding, Newborn Resuscitation Program, quality improvement program (Liu et al.), neonatal nutrition program, and respiratory support program (Bang, Bang, Baitule, Reddy, & Deshmukh, 1999; Baqui et al., 2008; Brotherton et al., 2021; Darmstadt et al., 2005; El-Atawi, Elhalik, & Dash, 2019; Koyamaibole, Kado, Qovu, Colquhoun, & Duke, 2006; Patel, Khatib, Kurhe, Bhargava, & Bang, 2017; Ramasethu, 2017; Shane & Stoll, 2014). Additionally, Every Newborn Action Plan (ENAP), was also established to reduce newborn deaths globally (Budetti & McManus, 1982; UNICEF, 2021b). Through collaboration mechanisms, ENAP made a concerted effort globally to reduce newborn deaths from 5 million to 2.4 million(W. UNICEF, 2021). Furthermore, the World Health Organization (WHO) is working closely with the ministries of health-Uganda and partners to strengthen and invest in healthcare service delivery, improve the quality of maternal and newborn care, expand the quality of healthcare service, reduce inequalities in healthcare services delivery, promote health infrastructures, promote community engagement, program tracking, and encourage accountability, with the intent of promoting newborn survival(WHO, 2021). Although several programs have been developed and put in place to prevent and reduce neonatal mortality, their significance is still inexplicable, and this failure contributed to the lack of Monitoring and evaluation systems in the day-to-day NICU clinical activities(Micah & Luketero, 2017). The current literature suggests that, integration of the Monitoring and Evaluation system components into the NICU’s activities will improve survival and reduce neonatal mortality rate (Mohamud, 2023; I. M. Njeru & Luketero, 2018; Ooko, Rambo, & Osogo, 2018).

In Uganda, neonatal mortality is a significant public health challenge, and these deaths are preventable in nature. Despite the availability of neonatal technological advancements in the country, the NICU’s performance in reducing and preventing newborn deaths is unsatisfactory. Many studies have tested the effectiveness of NICU after the implementation of; maternal and prenatal death reviews and surveillance (MPDRS)(Walugembe et al., 2024), weekly death surveillance, Kangaroo mother care(W. I. K. S. Group, 2021), Exclusive breastfeeding(N. S. Group, 2016), Newborn Resuscitation Program, quality improvement program, neonatal nutrition program, respiratory support program(Dol et al., 2018). Unfortunately, little is done to explore the performance and effectiveness of NICUs when integrated with Monitoring and Evaluation system components. The integration of Monitoring and Evaluation systems within the NICU is hypothesized to enhance the quality of care and reduce neonatal mortality by identifying and addressing gaps in the NICU service delivery. However, the incorporation of M&E systems into NICU is lacking in the health facilities operating in low resource setting raising concerns about achieving the global neonatal reduction target. There are 12 core components of M&E systems (Kevany et al., 2012; Ogungbemi et al., 2012) which are supposed to be institutionalized and incorporated into NICU’s daily activities which include; (1) Organization structures with M&E, (2) human capacity for M&E, (3) M&E partnership, (4) Costed M&E work plan, (5) M&E plan, (6) M&E advocacy, communication and culture, (7) Routine programming and monitoring, (8) Survey and surveillance (9) Data dissemination and use, (10) supervision and data auditing, (11) Evaluation and research, and (12) M&E databases (“UNAIDS,” 2009). However, based on the available resources only 4 (Four) M&E systems core components were studied include: (1) Organizational structures, (2) Human Capacity for M&E, (3) Routine Monitoring of NICU and (4) Supportive supervision and Data Auditing as described in **Figure 1** below.

**Figure 1.**
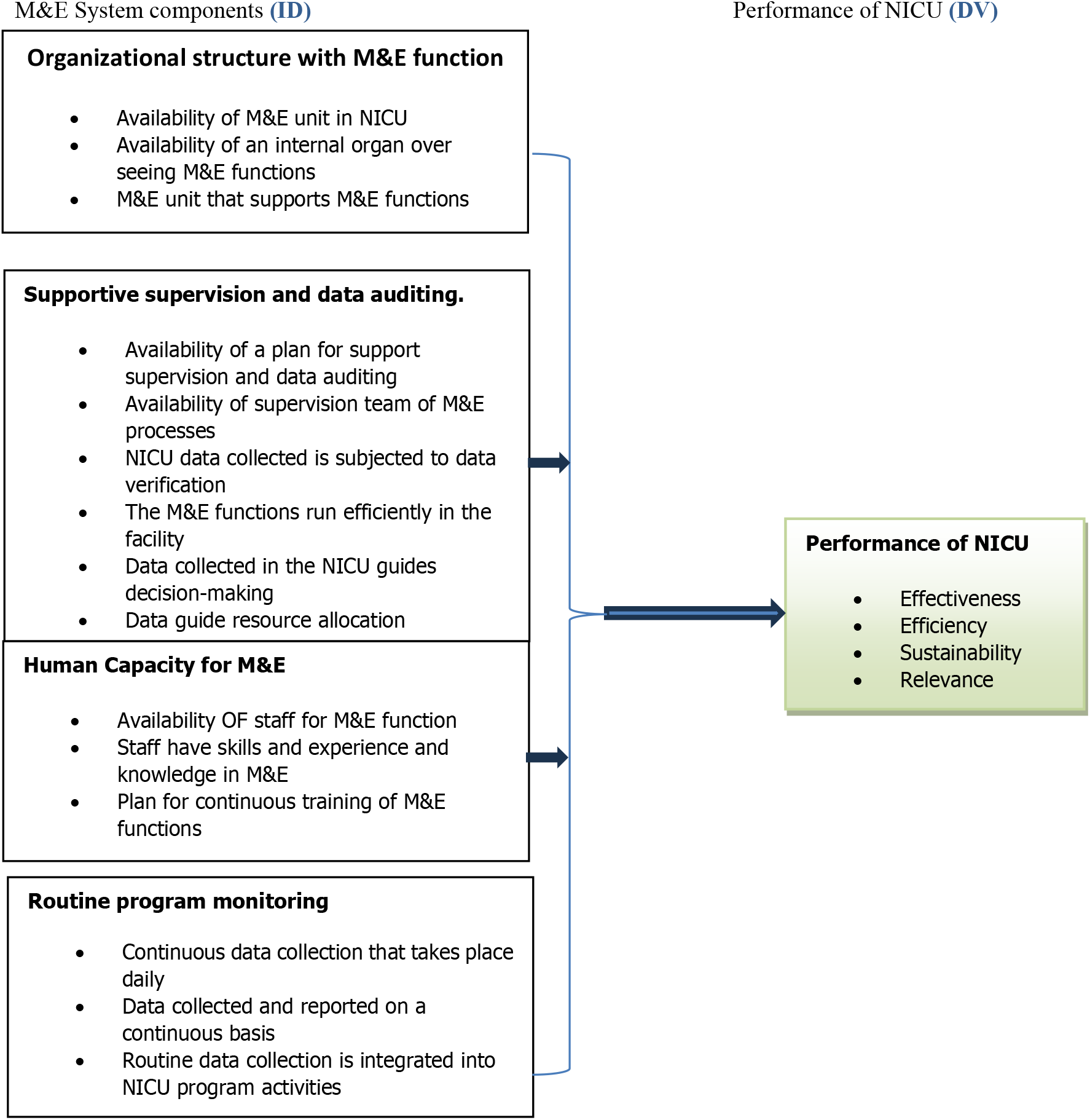
The conceptual framework illustrating the evaluated selected Monitoring and evaluation system components(Richard & Richard, 2019) in NICU performance.

Figure 1. *The conceptor framework indicates that;* Dependent Variable **“**Performance of NICU”. The overall effectiveness, efficiency, sustainability, and relevance of the NICU in providing neonatal care and reducing mortality rates. (a) Effectiveness: Was measured by the outcomes achieved, such as reduction in neonatal mortality rates. reduced length of hospitalization, the survival rate and improvement in healthcare indicators. (b) Efficiency: Evaluated by assessing the resources used relative to the outcomes achieved, including time, cost, and personnel. (c) Sustainability: Determined by the ability of the NICU to maintain its performance and outcomes over time without external support. And (d) Relevance: Assessed by the extent to which the NICU’s services meet the needs of the target population, including improving the quality of life of the newborns, reduced morbidity, responsiveness to changes in demand and healthcare needs.

Figure 1. Furthermore, indicates that, response variables include: (1) The response variables include: Organizational Structure with M&E Function; This variable refers to the formal framework within the NICU that supports and oversees the Monitoring and Evaluation (M&E) processes, and it was measured as availability of M&E unit in NICU, and it was assessed by checking whether a dedicated M&E unit exists within the NICU. Secondly, the availability of an internal organ overseeing M&E functions. This was determined by identifying any internal body or committee responsible for supervising M&E activities. Lastly, M&E unit that supports M&E functions evaluation was based on the level of support provided by the M&E unit to the NICU, including resource allocation, training, and supervision(Richard & Richard, 2019). (1) Supportive Supervision and Data Auditing. This variable pertains to the mechanisms in place to ensure that M&E processes are being followed and that data collected is accurate and reliable. (a) It was measured by assessing the availability of a plan for support supervision and data auditing. This was measured by the existence of documented plans outlining supervision and data auditing procedures. (b) Availability of a supervision team for M&E processes: This was evaluated by identifying whether a team is in place to oversee M&E activities. (c) NICU data collected is subjected to data verification: This was assessed by reviewing whether data collected from the NICU is routinely verified for accuracy. (d)The M&E functions run efficiently in the facility: it was determined by evaluating the efficiency of M&E processes, including timeliness and resource utilization. (e) Data collected in the NICU guides decision-making: This was assessed by analyzing how frequently data is used in making clinical and operational decisions. (f) Data guide resource allocation: This was evaluated by determining whether data-driven insights are used to allocate resources effectively within the NICU(Richard & Richard, 2019). (2) Human Capacity for M&E: Definition: This variable represents the skills, knowledge, and availability of personnel responsible for carrying out M&E functions within the NICU. Measurement was done based on the availability of staff for M&E function. This was determined by the presence of dedicated M&E staff within the NICU. (b) Staff have skills, experience, and knowledge in M&E, and this was evaluated by assessing the qualifications and experience of the staff involved in M&E activities. (c) Plan for continuous training of M&E functions, was determined by the existence of a training plan aimed at enhancing M&E skills among staff(Richard & Richard, 2019). (3) Routine Program Monitoring: This variable was looked at as a continuous collection, analysis, and reporting of data within the NICU to monitor its performance and outcomes. (a) Continuous data collection that takes place daily: Assessed by reviewing the frequency and regularity of data collection in the NICU. (c) Data collected and reported on a continuous basis: Evaluated by checking whether data is consistently reported to relevant stakeholders. (d) Routine data collection is integrated into NICU program activities: Determined by assessing how well data collection is embedded into the daily operations and activities of the NICU(Richard & Richard, 2019)

Although various health interventions like Maternal and Perinatal Death Review and Surveillance (MPDRS), Local Maternity and Neonatal care systems (LMNS), Kangaroo mother care (KMC), newborn resuscitation program, Weekly death review and surveillance have been implemented and studied within Neonatal Intensive Care Unit (NICUs), there is limited research comparing the performance of NICU with and without Monitoring and evaluation (M&E) systems.

This gap highlights the need for studies that specifically assess the impact of incorporating M&E systems on the effectiveness and outcomes of NICU services. The limited research that does exist often fails to provide a comprehensive analysis of how M&E components like routine program monitoring, human capacity for M&E, and organizational structures influence NICU outcomes. This gap underscores the need for more empirical studies to explore the potential benefits of M&E systems in enhancing NICU performance, particularly in resource-constrained settings. By comparing the NICU’s performance before and after the implementation of these M&E systems, the study seeks to provide empirical evidence on the role of M&E in improving neonatal care outcomes in the Yumbe region in Uganda.

## Methods

### Study design

The study used a separate-sample pretest-posttest quasi-experimental study design to compare the performance of the Neonatal Intensive Care Unit (NICU) with M&E systems(post-test) Versus NICU without monitoring and evaluation system(pre-test) in Yumbe region.

### Study participants

#### Eligibility criteria

The study recruited all the neonates admitted to the NICU at Yumbe Regional Referral Hospital, aged between 0(Zero) days to 27 days.

#### Recruitment process

Neonate-Mothers registered in the admission register book received an invitation letter from the researcher immediately after admission. This letter asked the neonate mothers whether they would like to complete a questionnaire survey about the performance of the NICU.

#### Recruitment setting

The recruitment of the study participants was conducted at the NICU of Yumbe Regional Referral Hospital. This was a Rolling recruitment process and it was done for the period of 7 months from (May 2023 to November 2023). The eligible participants who were available at the start of the study were also allocated to receive the intervention, and later a hard copy of the questionnaire surveys was hand delivered/ distributed to be completed by the mothers of infants in the NICU with the help of research assistants. This same process was repeated until the study obtained its required sample size. The participants were prompted to complete the survey within 5 to 10 minutes of receiving the survey, at time and allocation. Researcher assistants encouraged participants to complete the surveys in the allocation to which they belong.

### Intervention

#### Pretest

Before the intervention “Monitoring and Evaluation System”, NICU underwent an initial measurement (pretest) of its performance and effectiveness “Dependent variable”. The results established were the baseline for comparison.

The intervention “Monitoring and Evaluation System Components”, (Organisational structures, Human Capacity for M&E, Routine Monitoring of NICU and Supportive supervision and Data Auditing)(“UNAIDS,” 2009) were installed in the NICU and the selected components(were implemented for the duration of 105 days (One hundred and Five days) daily from Monday to Monday, at all times.

#### Posttest intervention

After the intervention (Monitoring and Evaluation system components), the performance of the NICU was measured again (posttest) using the dependent variable (Performance of NICU). The results obtained from the posttest were compared with those from the pretest.

#### Methods of Intervention Content Delivery

Monitoring and evaluation system components were installed in the NICU with an admission capacity of 30 neonates, although sometimes it admits between 20 -27 neonates, on average, representing 80% of its admission capacity. The components of the M&E systems allocated were; Monitoring and evaluation tools developed, and trained M&E personnel appointed to work in the NICU. Survey and surveillance, and data dissemination and use mechanism was put in the NICU. Human resources for monitoring and evaluation were appointed. Data collection tools were designed and used in the NICU. A computer and internet system were also installed in the NICU to store and analyze the data collected. All these components were delivered by trained M&E specialists and trained data persons. In the study, the frequency of exposure to the intervention was administered daily at all times for a duration of 105 (One hundred and five) days. The total duration set to run the intervention was 105 days including the total number of sessions/ exposures over the entire study period. The duration and frequency of the sessions were consistent throughout the intervention period with no variations in the schedule. To promote compliance, a peer support group was established, where participants shared experiences and encouraged each other, furthermore, the principal investigator was available for a one-on-one session to address any concerns of the participants. Lastly, progress tracking, accessibility to research materials, and engagement with stakeholders, incentives and rewards, sending reminders and follow-up techniques were used to increase adherence.

#### Specific objectives and hypothesis

The study aimed to explore the effect of Monitoring and Evaluation systems on the Neonatal intensive care unit (NICU) performance. The hypothesis was that the implementation of Monitoring and Evaluation systems will significantly improve the performance of the Neonatal intensive care Unit (NICU).

#### Outcomes

NICU performance was the primary outcome and it was assessed through its major performance key indicators (KPIs) which include; neonatal survival rates, readmission rates, morbidity rate, infection control, and incidences and complications. To measure the primary outcome (NICU performance), Data on these key performance indicators was collected before (pretest) and after(posttest) the implementation of the monitoring and Evaluation systems to compare and determine any significant change in NICU performance.

The secondary outcome was patient satisfaction. The degree of satisfaction among patients with the care provided in the NICU and it was assessed through standardized surveys and feedback forms distributed to the neonate mothers at both the pretest and posttest of the implementation of the monitoring and evaluation systems.

#### Measurement of NICU performance

The outcomes were assessed after collecting data from medical records, electronic health records, and NICU administrative databases. Data on the key performance indicators was extracted using the designed survey form, and it was collected weekly for a period of 7 seven months. The study used observation checklists, audit reports, and patient surveys as data sources. During the data collection process, the study conducted periodic audits and direct observations to assess adherence to protocols and administered surveys to NICU staff regarding their compliance and challenges. The observation and audits were conducted bi-weekly survey administered monthly.

#### Measurement of patient satisfaction

The study used standardized satisfaction surveys, feedback forms, and interviews with the neonate mothers. The data collection process was done by distributing surveys and feedback forms to NICU patients, furthermore, follow-up interviews were conducted to gather more in-depth feedback. The survey and feedback form distribution were done upon the patient’s discharge, and the interviews were conducted within one-week post-discharge. To enhance the quality of measurement; research assistants were trained on the study protocols, data collection tools, and ethical considerations. This minimized variability and errors in data collection. Secondly, the study used standardized and validated tools for data collection, such as established performance key indicators, validated survey instruments, and a reliable observation checklist. This ensured consistency and accuracy in the data collection. Additionally, the data collection tools were piloted. Inter-rater reliability checks for observational data were conducted to assess the inter-rater reliability by using multiple observers independently to assess the same events to ensure consistency in the observation. Lastly, data quality audits were conducted, blinding the data collectors, and analysis to the study hypothesis and intervention status of participants to reduce bias in the data collection and analysis.

#### Validity and reliability

The questionnaire survey and observation checklist were tested for validity and reliability by Calculating Cronbach’s alpha for the items in the questionnaire, a Value of 0.85 was obtained, suggesting the reliability level was acceptable (Wadkar, Singh, Chakravarty, & Argade, 2016). Pre-data collection, the instrument tools were administered to the participants to collect observational data at two different points, to calculate the correlation between the two sets of responses. For the observational checklist, an inter-rater Reliability scale was computed by employing multiple observers to rate the same response or behaviors and calculated the Inter-Rater reliability using Cohen’s Kappa, and interclass correlation coefficient as illustrated by Bi and Friends (ICC)(Bi & Kuesten, 2012).

#### Sample size estimation

In this pretest-posttest quasi-experiment study, the sample size was calculated using Krejcie & Morgan’s table. The study had a population **size of (N=600)**, and the table recommends a sample **size of 234** to achieve a 95% confidence interval level with a 5% margin of error. However, we increased the **sample size to 236(pretest group n=106, Posttest group n=130)**, participants to increase the robustness and account for potential dropouts. In this study, a three-stage interim analysis was conducted at the 25%, 50%, and 75% stages of the data collection period. The analyses conducted at different stages aimed to evaluate the monitoring and evaluation systems on neonatal mortality rates and to ensure that the implementation was proceeding as planned. At different levels, adjustments were made based on these interim findings to address any issues and optimize the effectiveness of the intervention.

### Methods of Assignment of the Intervention

#### Unit of assignment

In this study, the unit of assignment was at individual level, neonates and their mothers were admitted to the NICU ward at Yumbe Regional Referral Hospital. Neonates were individually assigned to either the pretest or posttest group based on their admission time relative to the implementation of the monitoring and evaluation system components. The method of assignment was: The Pretest group assignment had 103 neonates admitted to the NICU before the implementation of monitoring and evaluation systems. Eligible neonates admitted to the NICU between MAY 1, 2023, and August 31, 2023, were assigned to the pretest group. These neonates did not experience the newly implemented monitoring and evaluation systems. 130 neonates were assigned to the Post-test group. Eligible neonates were those admitted to the NICU after the implementation of the monitoring and evaluation systems components, between SEPTEMBER 1, 2023, and NOVEMBER 31, 2023. These neonates were subjected to the newly implemented monitoring and evaluation systems. The implementation process was; that the monitoring and evaluation system components were introduced in the NICU on SEPTEMBER 1, 2023. Neonates admitted before this date were included in the pretest group, while those admitted on or after this data were included in the post-test group. Data on neonatal mortality rates were collected for both groups to assess the effect of the intervention.

#### Blinding

In this study, blinding was made at three levels. Data collators were only provided with a unique identifier for each neonate, and they did not have access to information about when the neonate was admitted or whether they were subjects to the intervention. The outcome assessors were given mortality data without any timestamps or group labels, ensuring that their analysis was unbiased, of any knowledge of the intervention timing. The healthcare providers trained in the M&E systems continued their usual care practices without any additional information about the study’s timeline or group assignment.

#### Unit of Analysis

In this pretest-posttest study, the individual neonate was the primary unit of analysis. Data on each neonate, including mortality status, length of stay, complications, and demographic information, were collected and analyzed to assess the effects of the monitoring and evaluation system components on neonatal mortality rates.

#### Data analysis

For the descriptive statistics, an independent samples t-test, and Fisher’s exact test were computed to summarize the demographic and baseline characteristics of the participants. Measures of central tendency (mean, median) and dispersion (standard deviation, interquartile range) were calculated for continuous variables such as birth weight and length of stay. Frequency distributions and percentages were used to summarize categorical variables such as gender and mortality status. Inferential statistics were used to compare neonatal mortality rates between the pretest and post-test groups using chi-square tests and t-tests. The correlated data was assessed using generalized estimating equations and mixed-effects models. A subgroup and adjusted analyses were performed using stratified analyses and multivariate regression models, and means were compared using independent samples t-tests and effect size calculations. Bias was controlled using Logistic regression analysis to adjust for potential confounders, ensuring that the observed effects of the intervention on neonatal mortality were not due to these variables. To ensure validity and reliability; factor analysis was done, and reliability was computed at three levels; internal consistency was evaluated using Cronbach’s alpha, and it was (α = 0.86). Test-Retest Reliability was assessed with a coefficient of 0.82, and the Inter-Rater Reliability had a coefficient of 0.78. To test the normality of the data, the Shapiro-Wilk test was applied to the distribution of birth weight and length of stay. Since the p-values were greater than 0.05, the data were considered normally distributed. The best-fit model was selected based on AIC, BIC, likelihood ratio tests, R-squared values, Hosmer-Leme show tests, and residual analysis. Superiority and non-inferiority assessments were conducted to evaluate the intervention’s effectiveness. Missing data were handled using multiple imputation methods to minimize bias. The statistical analyses were conducted using software programs STATA (Release 17. College Station, TX: StataCorp LLC; 2021.).

#### Ethical Statement

Ethics approval was obtained for this study. The Uganda National Council for Science and Technology (UNCST) approved the research project under registration number HS497ES, with the approval valid from 13/04/2023 to 13/04/2024. Additionally, ethical clearance was granted by the Uganda Technology and Management University (UTAMU) on 03/04/2023. Informed Consent: Participants in the study were newborns, and therefore, parental / guardian consent was obtained in written form prior to participation. This consent process was properly documented and witnessed, in line with the ethical requirements

## Results

The study involved 236 Neonates, with 103 in the pre-test group and 130 in the Post-test group. Table 1. Summarise the demographic and baseline characteristics of the study participants

**Table 1.** Demographic characteristics of the study participants.

| Variables | <u>Pre-test group(n=103)</u> | <u>Post-test group(n=130)</u> | <u>Total (n=236)</u> |
| --- | --- | --- | --- |
| **Gender** |  |  |  |
| Male | 58(56.3%) | 70(53.8%) | 128(54.2%) |
| Female | 45(43.7%) | 60(46.2%) | 108(44.5%) |
| **Mean Birth weight (g)** | 2500 ± 500 | 2550 ± 450 | 2525 ± 475 |
| **Mean Gestational age (weeks)** | 37.5 ± 2.5 | 37.8 ± 2.3 | 37.6 ± 2.4 |
| **Mean length of stay(days)** | 10 ± 4 | 8 ± 3 | 9 ± 3,5 |
| **Neonatal Mortality** |  |  |  |
| Yes | 20(19.4%) | 10(7.7%) | 30(12.7%) |
| No | 83(80.6%) | 120(92.3%) | 203(87.3%) |

*The values are presented as Mean ± standard deviation (SD) for continuous variables, and Number (Percentages) for categorical variables*

**Table1**. The gender distribution was relatively balanced across both groups, with males comprising 56.3% of the pre-test group and 53.8% of the post-test group. The mean birth weight in the pre-test group was 2500 grams (SD=500), while in the post-test group, it was slightly higher at 2550 grams (SD=450). The mean gestational age was similar between groups, with the pre-test group averaging 37.5 weeks (SD=2.5) and the post-test group averaging 37.8 weeks (SD=2.3). The mean length of stay in the NICU was 10 days (SD=4) for the pretest group and 8 days (SD = 3) for the post-test group, indicating a reduction in hospital stay duration after the intervention. Neonatal mortality rates were significantly lower in the posttest group, with 7.7% mortality compared to 19.4% in the pretest group. The implication of these findings underscores the importance and impact of monitoring and evaluation systems intervention on neonatal outcomes, as reflected by reduced length of stay and lower mortality rates in the post-test group.

**Table 2.**
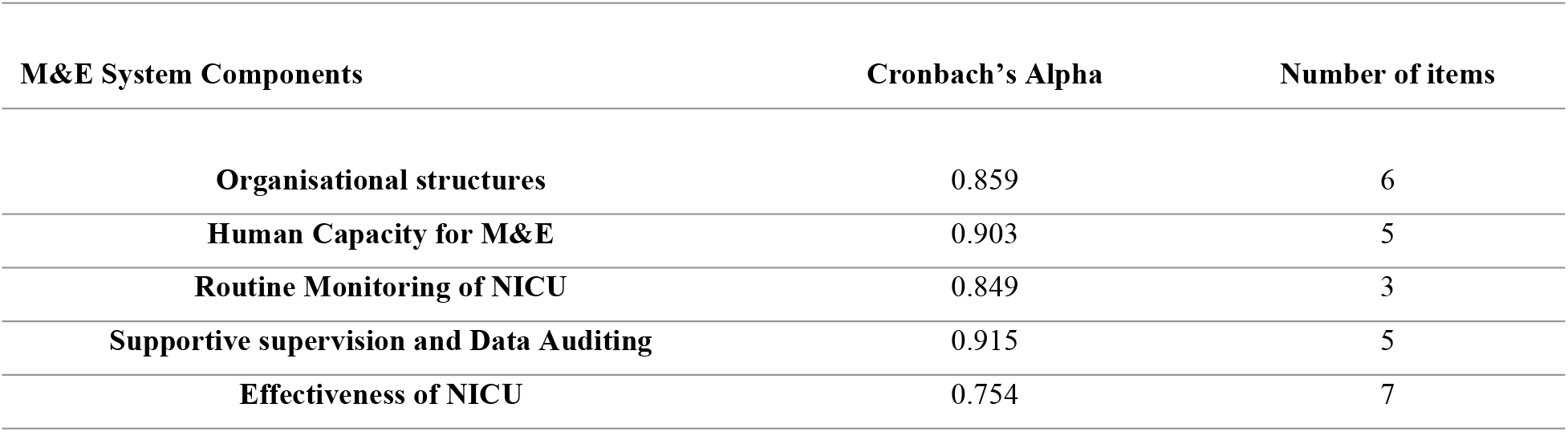
Reliability of likert Scale Data.

| M&E System Components | Cronbach's Alpha | Number of items |
| --- | --- | --- |
| Organisational structures | 0.859 | 6 |
| Human Capacity for M&E | 0.903 | 5 |
| Routine Monitoring of NICU | 0.849 | 3 |
| Supportive supervision and Data Auditing | 0.915 | 5 |
| Effectiveness of NICU | 0.754 | 7 |

The Cronbach’s alpha values for all M&E system components range from 0.754 to 0.915, indicating that the Likert scale items used in the questionnaire have excellent internal consistency. This supports the reliability of the instrument used to measure various components of the Monitoring and evaluation systems in the NICU.

**Table 3**. The effectiveness of the neonatal intensive care unit (NICU) before the intervention had a weighted average mean of 2.8, while in the post-test group improved to 3.5(p< 0.01), indicating a significant positive change following the intervention. The Organizational structures with the M&E function had a mean response of 2.0 in the pre-test group and improved to 2.5(p= 0.03) demonstrating a statistically significant improvement. The supportive supervision and data audit had increased mean response from 3.0 in the pre-test group to 3.5(p=0.04) in the post-test group, which indicates a significant improvement. Routine monitoring of NICU activities in the pre-test group had a weighted average mean of 2.9, while in the post-test group, it was 3.3(p=0.06) this indicates a trend towards improvement but not reaching conventional significance, while human capacity for M&E had a significant increase in the mean response in the post-test group of 3.2(p=0.02) higher than that of pre-test group of 2.5 weighted average mean response. Overall, the findings illustrate a clear and significant improvement in the post-test group, reflecting the effectiveness of the M&E systems intervention towards improving the performance of the NICU.

**Table 3.** Weighted average mean of M&E system components.

| M&E systems | Pre-test group mean | Post-test group mean | Weighted Average mean | P-Value Comparison |
| --- | --- | --- | --- | --- |
| Effectiveness of NICU | 2.8 | 3.5 | 3.3 | <0.01 |
| Organizational structure with M&E function | 2.0 | 2.5 | 2.42 | 0.03 |
| Supportive supervision and Data auditing | 3.0 | 3.5 | 3.42 | 0.04 |
| Routine monitoring of NICU activities | 2.9 | 3.3 | 3.18 | 0.06 |
| Human capacity for the M&E | 2.7 | 2.9 | 2.78 | 0.12 |
| *Weighted Average mean values reflect the mean response across all participants. P-values indicate the significance of the difference between pre-test and post-test groups, calculated using independent samples t-test* |  |  |  |  |

**Table 4**. Results from logistic regression analysis revealed that better organisational structures with M&E functions (OR=1.30, 95% CI: 1.10 – 1.55, P= 0.02) and supportive supervision (OR= 1.45, 95% CI: 1.15 – 1.80, P =0.01) significantly improve NICU effectiveness. Additionally, having adequate human capacity for M&E (OR = 1.35, 95%, CI: 1.05 – 1.75, P= 0.04) is a crucial factor for enhancing NICU performance. While routine monitoring of NICU activities shows a positive trend (OR= 1.20, 95% CI: 0.95 – 1.50, P= 0.10) it does not reach statistical significance, indicating the need for further investigation. These findings underscore the critical role of establishing structured M&E systems in the NICU, additionally, effective supervision and data audit, and sufficient human resources for M&E are crucial in improving the quality and outcomes of neonatal care in the NICU.

**Table 4.** The Impact of M&E systems on NICU performance.

| Variable | Odds Ratio<br>(OR) | 95% Confidence<br>Interval | P- Value |
| --- | --- | --- | --- |
| **organizational structure with M&E function** | 1.30 | [1.10 –1.55] | 0.02* |
| **Supportive supervision and Data audit** | 1.45 | [1.15 –1.80] | <0.01* |
| **Routine monitoring of NICU activities** | 1.20 | [0.95 –1.50] | 0.10** |
| **Human capacity for M&E** | 1.35 | [1.05-1.75] | 0.04* |
| ** Non-significant, * Significant p-value o P- value was obtained from logistic regression, CI= confidence interval, OR= odds ratio |  |  |  |

**Table 5**. The implementation of monitoring and evaluation (M&E) systems in the NICU led to significant improvements across multiple performance metrics. The effectiveness score of the NICU increased significantly from 2.75 in the pre-test phase to 3.30 in the post-test phase (ρ < 0.01), indicating a notable enhancement in overall NICU performance. Additionally, the average length of stay in the NICU decreased significantly from 12.0 days to 7.5 days(ρ=0.02), suggesting that the M&E systems intervention contributed to quicker recovery and discharge times for neonates. Staff compliance with NICU protocols also improved significantly, rising from 70% to 85% (ρ< 0.01) reflecting better adherence to clinical guidelines, likely due to the influence of M&E systems. Moreover, the neonatal mortality rate decreased significantly from 25% in the pre-test phase to 10% in the post-test phase (ρ< 0.05), underscoring the positive impact of the M&E systems on neonatal survival rate. These findings demonstrate the critical importance of M&E systems in optimizing neonatal care and improving outcomes, reinforcing their value in high-risk healthcare settings.

**Table 5.** The performance of NICU before and after the implementation of M&E systems.

| NICU Performance Matric | Pre-test Value | Post-test Values | Improvement | P-value |
| --- | --- | --- | --- | --- |
| NICU Effectiveness (Mean ) | 2.75 | 3.30 | +0.55 | < 0.01 |
| Length of stay(mean day) | 12.0 | 7.5 | -4.5 days | 0.02 |
| Staff Compliance (%) | 70% | 85% | +15% | < 0.01 |
| Neonatal mortality rate (%) | 25% | 10% | -5% | < 0.05 |

## Discussion

The study aimed to explore the effect of Monitoring and Evaluation (M&E) systems on the performance of Neonatal Intensive Care Unit (NICU) in the Yumbe region. The Implementation of the M&E systems significantly influenced several key performance indicators of NICU which include; Neonatal survival rates, neonatal mortality, and length of stay significantly improved. The study findings provide compelling evidence of the significant impact that monitoring and evaluation (M&E) systems can have on the performance of Neonatal Intensive care unit (NICU), particularly in law-resource setting such as Yumbe region. The results underscore the crucial role of M&E systems in enhancing NICU outcomes, including neonatal survival rates, reduced neonatal mortality, length of hospitalization, and overall performance. The introduction of M&E systems in the NICU led to notable improvements across several key performance indicators, which aligns with findings from previous research indicating that structured M&E frameworks can significantly enhance healthcare quality and outcomes (Kiboi, Kilonzo, & Iravo, 2018). The study’s primary finding-improved NICU performance with the implementation of M&E systems is consistent with the literature highlighting the effectiveness of M&E systems in optimizing newborn clinical care (Hamad & Ahmed, 2016; Kiboi et al., 2018; E. M. Njeru & Obwatho, 2018). The M&E systems not only improved neonatal survival rate but also led to shorter length of hospitalization and higher rotation rates, reinforcing the utility of real-time data and feedback in making informed newborn care clinical decisions. This aligns with studies demonstrating that advanced monitoring and evaluation technologies are associated with reduced advance events, improved surveillance system, hence better patient outcomes (Biwott, Egesah, & Ngeywo, 2017) (Kwast, 1998; Plevris & Lees, 2022). Among the specific components of the M&E systems, organizational structures with defined M&E functions, enhanced human capacity for M&E, and robust supportive supervision and data auditing emerged as particularly influential. These elements were crucial in fostering an environment conducive to continuous improvement and effective healthcare service delivery. The findings are consistent with research indicating that well-structured M&E frameworks and adequately trained staff are vital for optimizing healthcare performance (Biwott et al., 2017). The study also highlights the importance of routine program monitoring, which contributed to better NICU performance by ensuring ongoing assessment and adjustments to care protocols. This supports the recommendation that NICUs should integrate regular reviews and feedback mechanisms into their operations to sustain and improve performance. However, the study’s generalizability may be limited by its specific context. The findings are based on a single regional referral hospital in the low-resource-constrained setting which may not fully represent other healthcare facilities” environment “. Additionally. While the study shows strong associations between M&E systems and NICU performance, it does not establish direct causation and unmeasured factors could influence the results. The quality of data collection and potential biases in the data reporting could also affect the findings’ reliability

## Conclusion

This study underscores the critical role of M&E systems in improving NICU performance through reducing neonatal mortality and improve newborn survival. Investing in M&E infrastructure is essential for enhancing neonatal care, particularly in underserved regions. Hospital should prioritize the development and implementation of robust M&E systems, forcusing on improving organizational structures for M&E, training staff and ensuring effective supportive supervision and routine monitoring. Such investments are cruicial for optimizing healthcare delivery and achieving better outcomes for neonatres West-Nile region.

## Data Availability

All data supporting the findings of this study are available upon request. Data underlying the results presented in this manuscript can be accessed by contacting the corresponding author at Innocent Ssemanda, Uganda Technology and Management University, via email at. The data are stored in a secure database and can be provided in accordance with applicable ethical and legal regulations. For any inquiries or requests regarding data access, please reach out to the corresponding author.

## Acknowledgments

We express our gratitude to the staff of the Neonatal Intensive Care Unit (NICU) at Yumbe Regional Referral Hospital for their support and collaboration during this study. Special thanks go to the entire staff of Yumbe Regional Referral Hospital, the team at Cleveland Specialized Hospital, and colleagues at Uganda Technology and Management University for their valuable contributions. We also acknowledge the support from the Ministry of Health and the CDC in Uganda.

